# Diagnostic Accuracy of Preoperative CT Staging for Esophageal Cancer for Surgical Patients at Public Hospitals, Addis Ababa, Ethiopia

**DOI:** 10.1101/2025.08.01.25332671

**Authors:** Hanna Damtew Tadesse, Abigael Abiy Mesfin, Michael Azeze Negussie, Meskerem Dessie Demessa, Mikiyas Gifawossen Teferi, Samuel Zerihun Tesfaye

## Abstract

**Background:** Esophageal cancer is a major health concern in Ethiopia and is often diagnosed at advanced stages because of limited access to diagnostic tools. Computed tomography is the primary imaging modality used for preoperative staging at Tikur Anbessa Specialized Hospital and its affiliated centers. However, its diagnostic accuracy has not been well studied locally.

**Objective:** To evaluate the diagnostic accuracy of preoperative CT TNM staging of esophageal cancer in surgical patients, postoperative histopathological findings were used as the gold standard.

**Method:** This retrospective cross-sectional study included 121 patients with histologically confirmed esophageal cancer who underwent preoperative CT staging and surgery between January 2021 and December 2024 at TASH and affiliated hospitals. Data were collected from medical records and CT images and analyzed via SPSS version 27. The sensitivity, specificity, PPV, NPV, and diagnostic accuracy were calculated.

**Results:** Among the patients, 57.02% were female and 42.98% male, with a median age of 54 years. SCC was the predominant histologic type (84.3%). Preoperative CT staging revealed T3 in 67.8% and T4 in 32.2% of patients. Nodal staging revealed N0 in 77.7% of the patients. The diagnostic accuracies of CT for the T3 and T4 stages were 49.6% and 67.8%, respectively. For the N0 to N3 stages, the accuracy ranged from 61.9% to 95%. The combined sensitivity and specificity for T staging were 82.6% and 20%, respectively; for N staging, they were 88.1% and 25.4%, respectively.

**Conclusion:** CT imaging has moderate accuracy in staging esophageal cancer but has limitations, particularly in differentiating tumor depth and nodal involvement. These findings underscore the need for multimodal imaging approaches, including MRI, PET, and EUS, where available, to improve preoperative assessment and patient outcomes.

## Introduction

Esophageal cancer is a leading cause of cancer-related morbidity and mortality worldwide, with an estimated 572,034 new cases and 508,585 deaths reported globally in 2018 (1). It is the fifth most common cause of cancer-related death in men and the eighth leading cause in women(2). In Ethiopia, esophageal cancer is a significant public health concern, with a crude incidence rate of 2.4 per 100,000 people annually, particularly affecting rural populations in the southern and eastern regions(3). The two most common histological types are squamous cell carcinoma (SCC) and adenocarcinoma, accounting for more than 90% of cases(4). Accurate preoperative staging is critical for determining the appropriate treatment strategy, assessing resectability, and predicting patient prognosis(3).

CT is widely used as the initial imaging modality for the preoperative staging of esophageal cancer because of its ability to evaluate tumor size, local invasion, lymph node involvement, and distant metastasis(4). However, the diagnostic accuracy of CT varies depending on the tumor stage and the experience of the radiologist. While CT is effective in assessing advanced disease, its accuracy in distinguishing early T stages (T1, T2, and T3) and detecting nodal involvement remains suboptimal(5). Endoscopic ultrasound (EUS) and positron emission tomography (PET) are often used as complementary imaging modalities to improve staging accuracy, but their availability is limited in resource-constrained settings such as Ethiopia (6).

In Ethiopia, esophageal cancer patients often present at advanced stages due to limited access to early diagnostic tools and specialized care (7). Preoperative staging relies primarily on CT imaging, but its diagnostic accuracy in this population has not been thoroughly evaluated.

Studies from other regions have shown that CT scans can reveal understage or overstage esophageal cancer, leading to inappropriate treatment decisions(2). For example, the inability of CT to differentiate between inflammatory and malignant lymph nodes often results in overstaging, whereas its limited resolution for detecting micrometastases can lead to understaging (8). These inaccuracies can significantly impact patient outcomes, particularly in settings where advanced imaging modalities such as EUS and PET are not readily available.

At Tikur Anbessa Specialized Hospital (TASH) and its affiliated hospitals, CT is the primary imaging modality for the preoperative staging of esophageal cancer. However, there is a lack of local data on the diagnostic accuracy of CT staging compared with surgical and histopathological findings. This gap in knowledge hinders the optimization of treatment strategies and limits the ability to provide evidence-based recommendations for improving patient care. Therefore, there is an urgent need to assess the diagnostic accuracy of preoperative CT staging in this setting to guide clinical decision-making and improve patient outcomes.

Accurate staging is essential for determining resectability, planning surgical interventions, and selecting appropriate neoadjuvant or adjuvant therapies(3). In resource-limited settings such as Ethiopia, where advanced imaging modalities are scarce, CT remains the cornerstone of preoperative staging. However, its limitations in early T staging, nodal involvement, and distant metastasis detection necessitate a thorough evaluation of its diagnostic accuracy in this context (5).

This study aims to address the gap in local data by assessing the diagnostic accuracy of preoperative CT staging of esophageal cancer at TASH and its affiliated hospitals, with correlations with surgical and histopathological findings. This study aims to contribute to the global body of knowledge on esophageal cancer staging, particularly in low-resource settings, and will inform future research on the integration of complementary imaging modalities to improve diagnostic accuracy(6). Ultimately, this study seeks to improve patient outcomes by ensuring that preoperative staging is as accurate and reliable as possible. The objective of this study was to evaluate the diagnostic accuracy of preoperative CT staging of esophageal cancer in patients undergoing surgery, with correlations with surgical and histopathological findings.

## Methods

### Study Setting

The study was conducted at Tikur Anbessa Specialized Hospital (TASH) and its affiliated hospitals in Addis Ababa, Ethiopia. Established in 1974, TASH is the largest tertiary hospital in the country and is administered by Addis Ababa University.

The radiology department at TASH provides a wide range of imaging services, including approximately 840 chest imaging studies per month. The cardiothoracic imaging unit is staffed by three senior cardiothoracic radiologists and two cardiothoracic radiology fellows, who actively provide services. Additionally, the unit holds multidisciplinary team (MDT) meetings every other week, involving oncologists, cardiothoracic surgeons, and radiologists. During these meetings, a minimum of three esophageal cancer cases are discussed per session.

TASH’s affiliated hospitals, such as St. Peter Referral Hospital and Menelik II Hospital, also contributed to the study by expanding the scope of patient data and providing additional training opportunities for residents and fellows.

### Study population

The source population included all patients with confirmed esophageal cancer through endoscopic biopsy who underwent preoperative CT imaging and surgery at TASH and its affiliated hospitals between January 2021 and December 31, 2024.

### Study Design and Study Period

This study was a hospital-based, retrospective, cross-sectional study. It was conducted at TASH and its affiliated hospitals, covering the period from January 1, 2021 to December 31, 2024. Patients with confirmed esophageal cancer through endoscopic biopsy and preoperative CT imaging were included. The data was accessed on Oct 1, 2024. The authors had access to identifiable patient information during the initial phase of data collection. Subsequently, all data were made anonymous prior to data analysis to ensure confidentiality. The CT images at initial diagnosis were collected and analyzed for tumor size, location, presence of calcification, and enhancement patterns. Additionally, image-defined risk factors and the presence of metastasis were evaluated and recorded.

### Sample size determination

All patients with esophageal cancer who underwent preoperative CT staging and surgery at TASH during the specified study period (January 2021 to December 31, 2024) were enrolled in the study.

### Sampling Technique

A total population sampling technique was used; all patients who met the inclusion criteria during the study period were included.

### Inclusion and Exclusion Criteria

#### Inclusion criteria

- All patients with esophageal cancer underwent preoperative CT staging and subsequent surgery (including intraoperative staging).
- Patients who had preoperative chest CT staging performed at facilities outside TASH but underwent surgery at TASH or its affiliated hospitals.
- For patients with multiple preoperative CT stages, the most recent preoperative CT stage was used.

#### Exclusion criteria

- Patients with esophageal cancer who lacked preoperative CT staging reports or whose CT images were not evaluated by a radiologist.
- Patients who did not undergo surgery.
- Patients who received preoperative neoadjuvant therapy without prior staging.

### Data collection and management

Patient medical records were accessed via patient identification numbers through the hospital’s MedWeb system. The CT images of patients with endoscopy-proven esophageal cancer were identified and evaluated. Key data, including the site of the lesion, TNM stage, type of surgery performed, and postoperative biopsy results, were recorded in an Excel spreadsheet for further analysis. The primary investigator checked and ensured that the data were clean and complete. To ensure the accuracy and reliability of the data, the primary investigator conducted regular and periodic supervision during the data collection process.

### Data analysis

The collected data were coded, cleaned, and thoroughly examined for completeness. It was then entered into the Statistical Package for Social Sciences (SPSS) version 27 software for analysis. Descriptive statistics, including frequency distributions, percentages, means, and standard deviations, were used to characterize the variables. Diagnostic accuracy, positive predictive value, negative predictive value, sensitivity and sensitivity were calculated for the TNM stages.

### Ethical considerations

Ethical clearance for this study was obtained from the Radiology Department’s Research and Publications Committee of the School of Medicine, College of Health Sciences, and Addis Ababa University under reference number DRPC/004/2017. The confidentiality of patient information was strictly maintained throughout the data collection, analysis, and dissemination of the results. Oral informed consent was taken from all participants through telephone communication and recorded on a consent log. The participants confirmed understanding and voluntarily agreed to participate. The study did not include minors.

## Results

A total of 121 patients fulfilled the inclusion criteria. A total of 86 patients were from Tikur Anbessa Specialized Hospital (TASH), 12 patients were from Menelik II referral hospital, and 23 patients were from ST Peter hospital, as shown in Figure 1.

**Fig 1.**
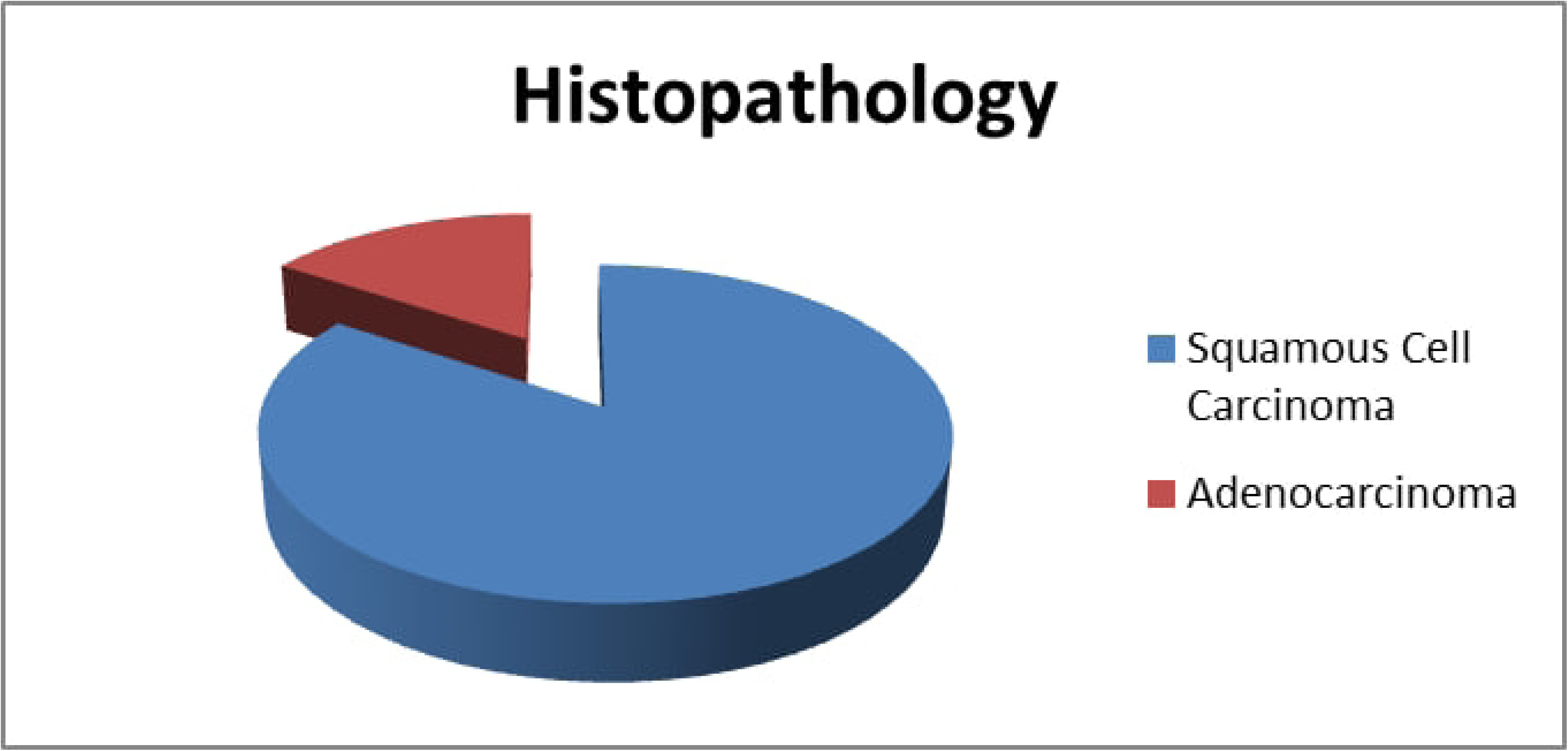
Patient Hospital Distribution

Among the study subjects, 69 (57.02%) were female, and 52 (42.98%) were male, as shown in Figure 2.

**Fig 2.**
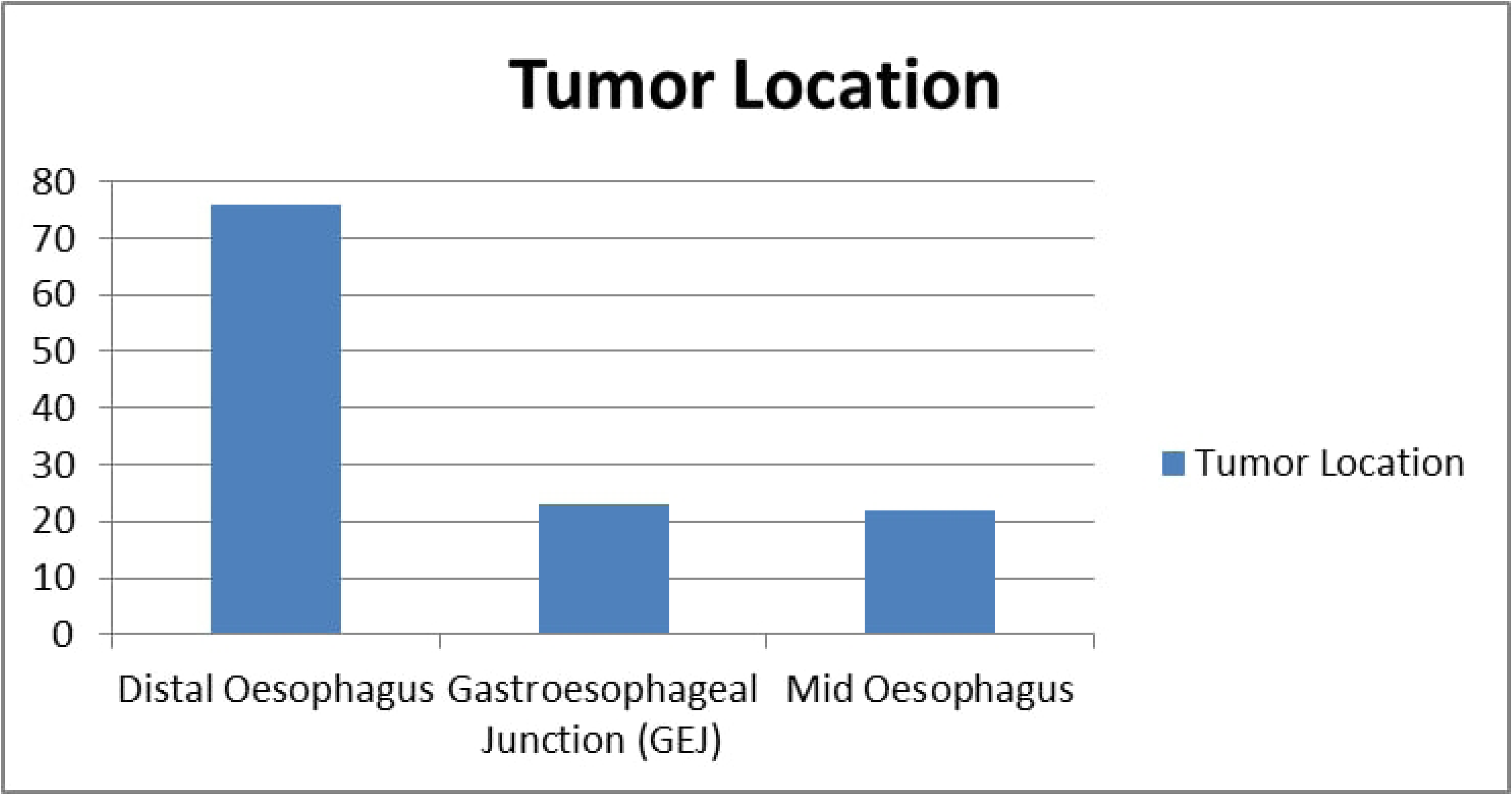
Patient Gender Distribution

The majority of patients were from the Oromia region (55%), followed by Addis Ababa (42%) and Amhara (9%), as described in Figure 3.

**Fig 3.**
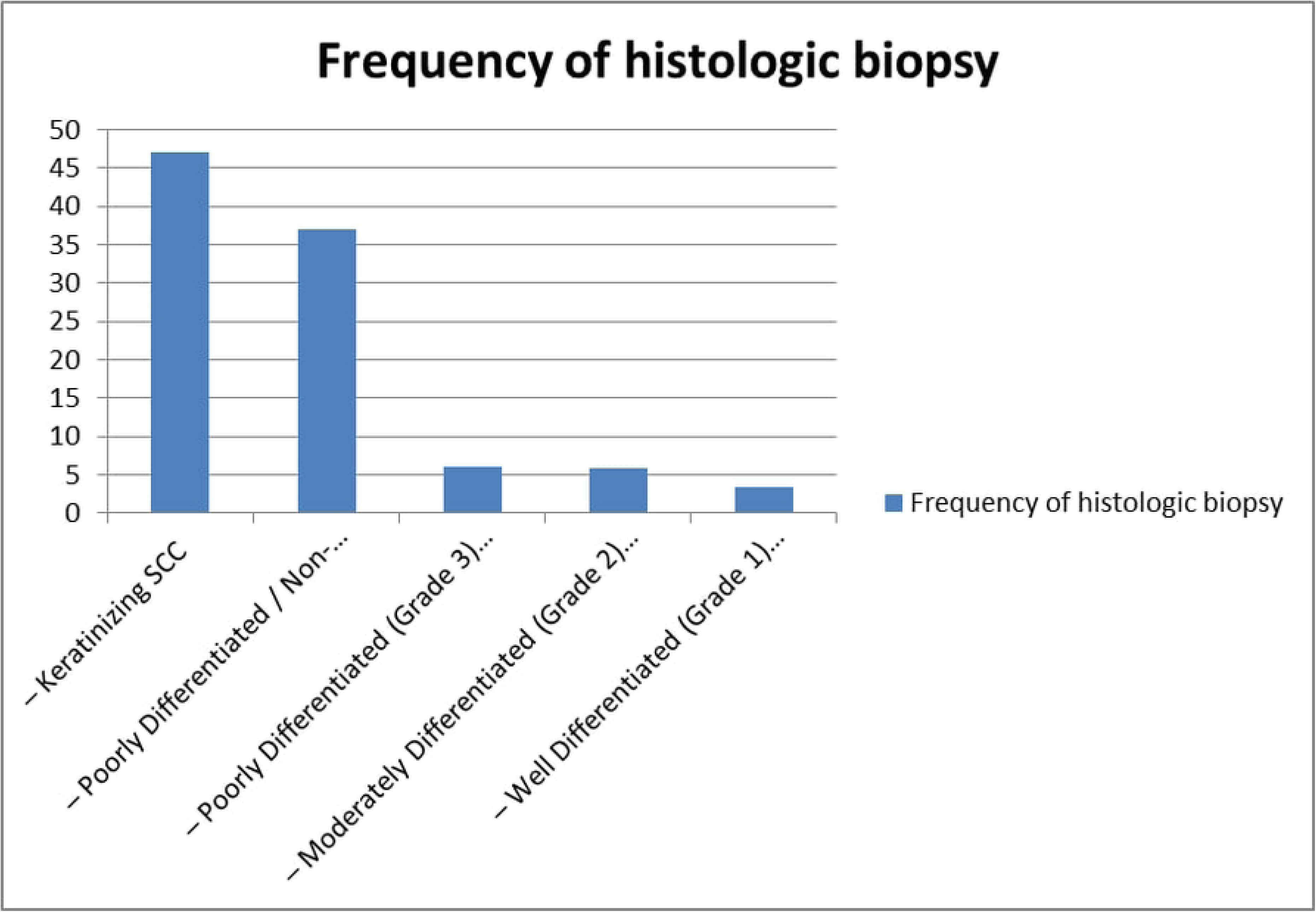
Regional distributions of patients

The age of the patients ranged from 16--91 years in the study, with a median age at presentation of 54. All patients had dysphagia at presentation, followed by vomiting, weight loss and vomiting as a frequent presentation. Among the 121 patients, only 3 patients (2.5%) mentioned that they had a past history of alcohol intake, and 4 (3.3%) patients mentioned having a past history of smoking. The average time interval between the preoperative CT scan date and the surgery date was 27.3 ± 11.1 days. According to the postsurgical histopathology results, 102 (84.3%) patients had confirmed SCC, and 19 (15.7%) patients had adenocarcinoma, as shown in Figure 4.

**Figure 4.**
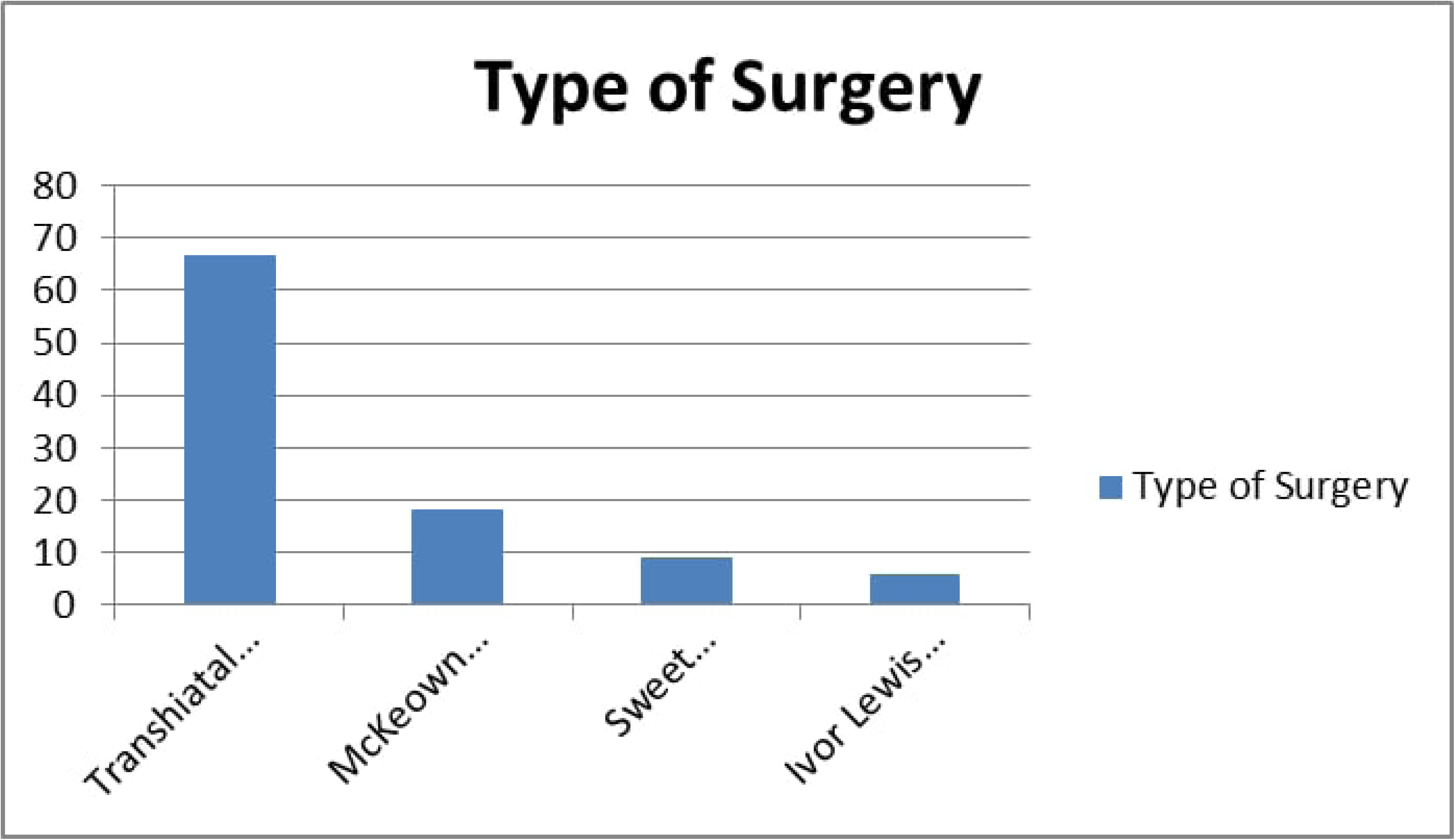
Histopathology results of patients

Endoscopic biopsy with histologic grading and differentiation revealed that the location of the mass at presentation was the distal esophagus in 76 (62.8%) patients, the GEJ in 23 (19%) patients and the middle esophagus in 22 (18.2%) patients, as shown in Figure 5. Among the patients who presented with SCC, the majority (47.1%) had keratinizing squamous cell carcinoma, whereas 37.1% of the patients had poorly differentiated SCC or nonkeratinizing SCC. As shown in Figure 6, among patients who were diagnosed with the adenocarcinoma type of esophageal cancer, poorly differentiated or Grade 3 adenocarcinoma was the most common histologic finding, accounting for 6.1% of the study participants.

**Figure 5.**
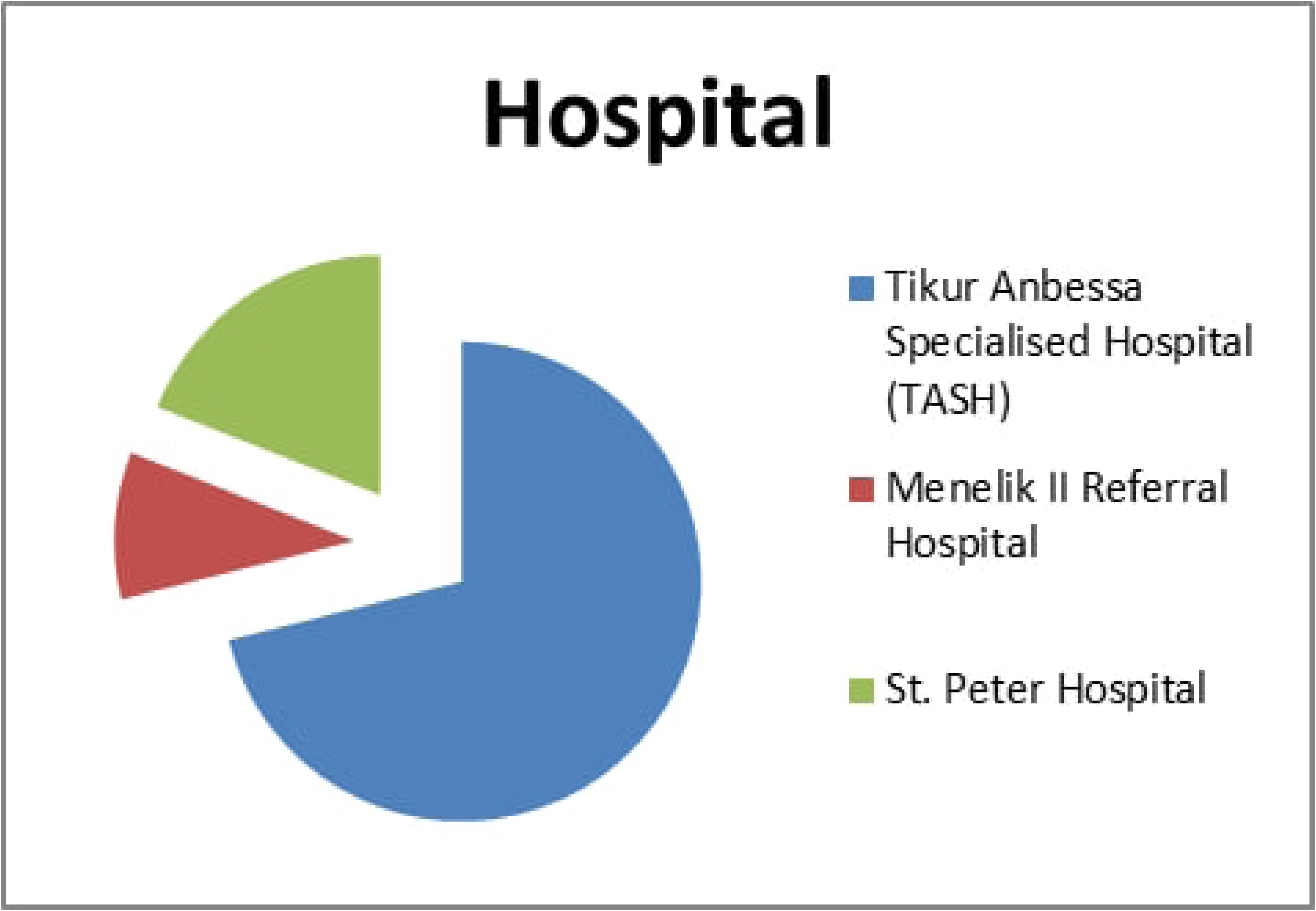
Frequency of tumor location

**Figure 6.**
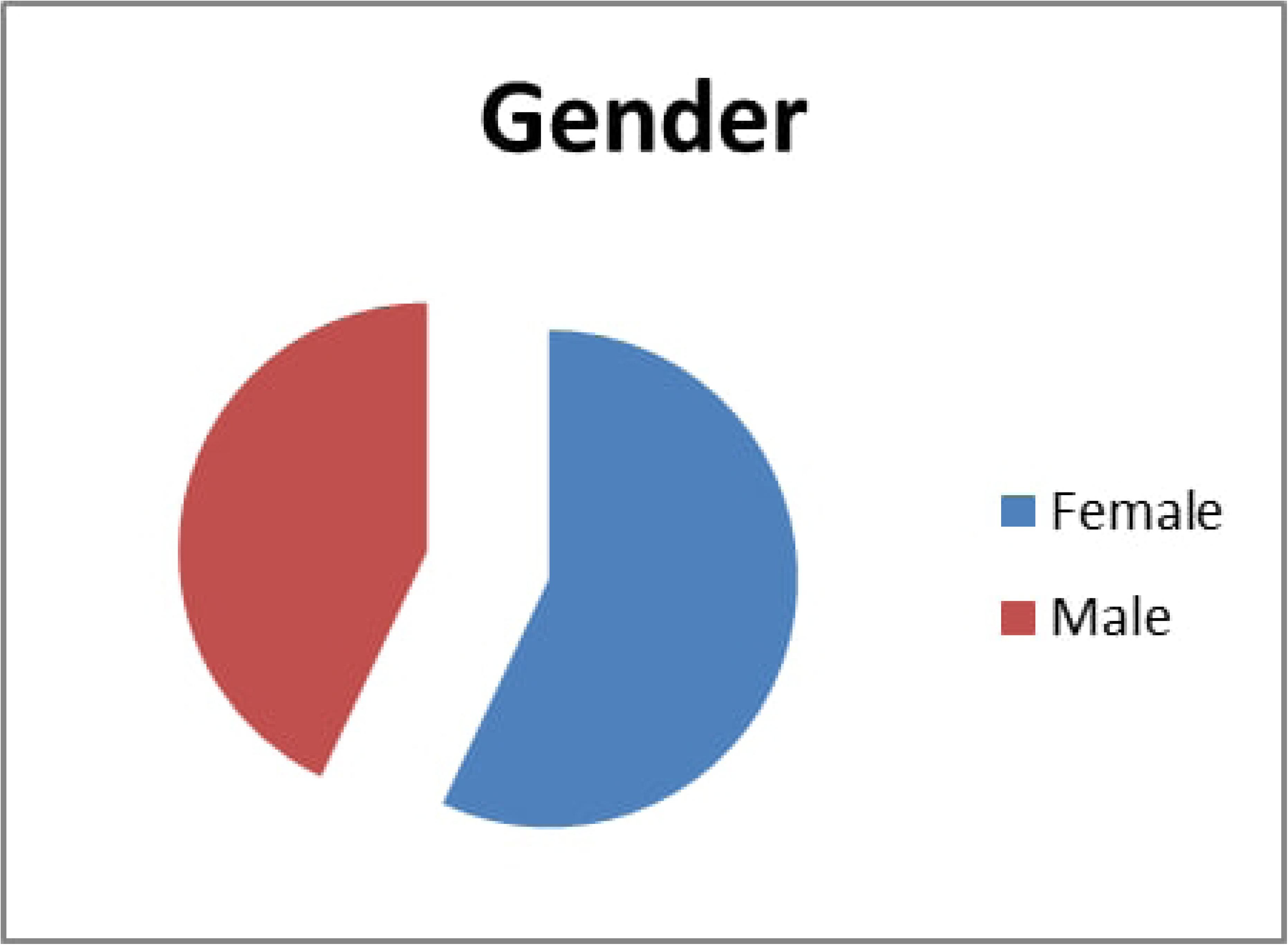
Frequency of histologic biopsy

On endoscopic evaluation of esophageal cancer, 47 (38.8%) patients had masses located 36–40 cm from the incisor, and 38 (31.4%) patients had masses 31–35 cm from the incisor, as described in Figure 7.

**Figure 7.**
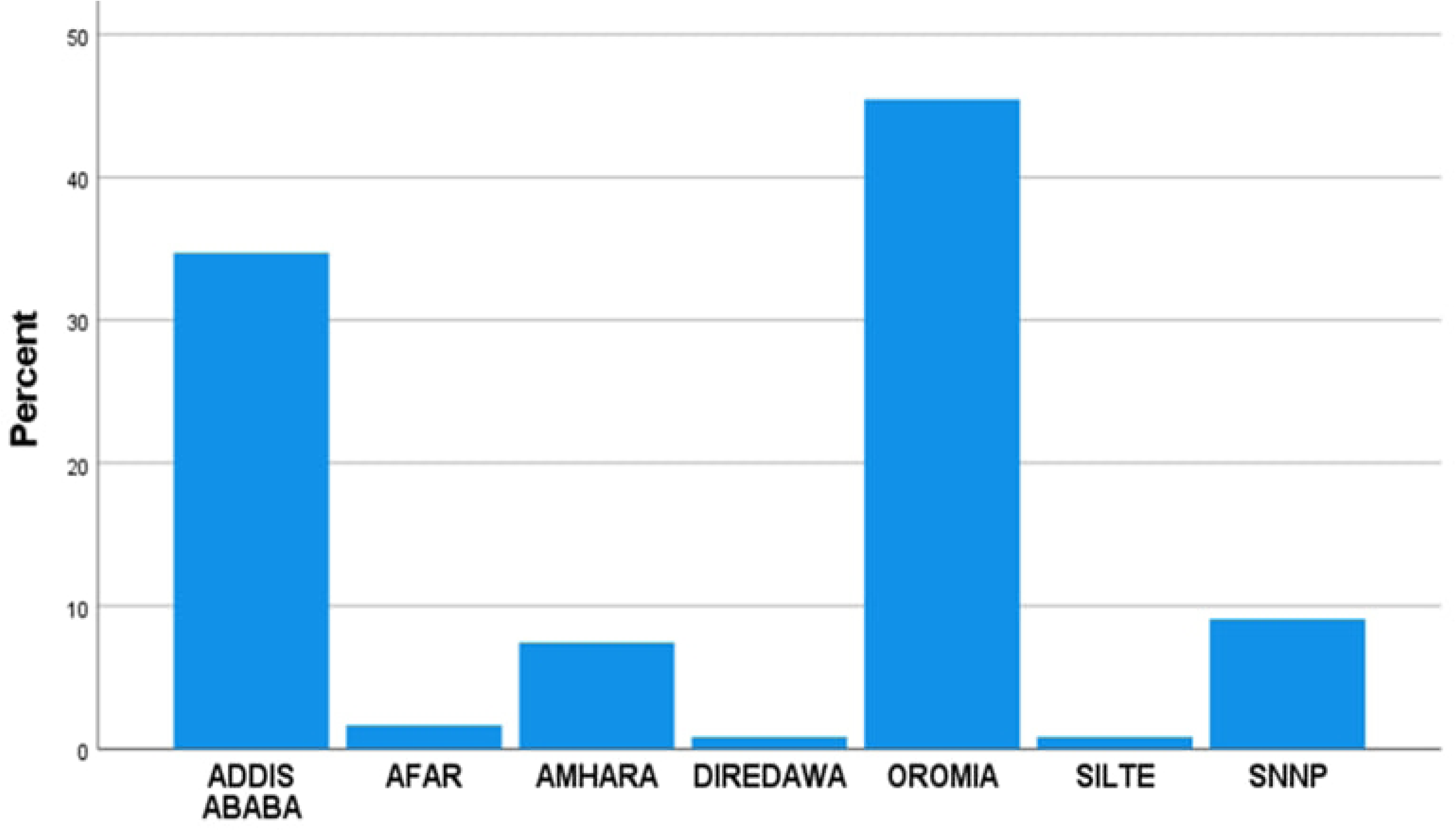
Types of surgery performed.

Among 121 patients with confirmed esophageal cancer, 82 (67.8%) had T3 disease, and 39 (32.2%) had stage T4 disease, as shown in Table 1. With respect to nodal diagnosis, 94 (77.7%) patients had no nodal involvement (N0) at presentation, and 15 (12.4%), 9 (7.4%) and 3 (2.5%) patients had N1, N2 and N3 disease at presentation, respectively. All patients had no records of distant metastasis.

**Table 1.** Distribution of preoperative esophageal cancer TNM CT staging at presentation.

| Stage |  | Frequency (n) | Percentage (%) |
| --- | --- | --- | --- |
| T Stage | T3 | 82 | 67.8 |
|  | T4 | 39 | 32.2 |
| N Stage | N0 | 94 | 77.7 |
|  | N1 | 15 | 12.4 |
|  | N2 | 9 | 7.4 |
|  | N3 | 3 | 2.5 |
| M Stage | M0 | 121 | 100.0 |

The size of the esophageal mass was measured as the total horizontal diameter of the mass ranged from 0.8 cm to 6 cm, with a mean diameter of 1.8 cm, and the total vertical (craniocaudal) diameter of the esophageal mass ranged from 2 cm to 9 cm, with a mean diameter of 5.5 cm. A total of 113 (93.4%) patients had a total vertical length greater than or equal to 3 cm. Eighty-one patients (66.9%) underwent transhiatal esophagectomy (THE), followed by McKeown esophagectomy, in 22 (18.2%) patients, as described in Figure 7.

Postsurgical histopathology biopsy results revealed that 69 patients (57%) had stage PT3 disease, 42 patients (34.7%) had stage PT2 disease, 5 patients (4.1%) had stage PT4A disease, and 5 (4.1%) had stage PT4B disease. Among the 121 patents who underwent OE for confirmed esophageal cancer, 70 patients (57.9%) had no nodal involvement (PN0), 30 patients (24.8%) had pathologic N1 disease, 18 patients (14.9%) had pathologic N2 disease, and only 3 patients (2.5%) had pathologic N3 disease, as shown in Table 2.

**Table 2.** Distribution of postoperative pTNM stages.

| Pathologic Stage | Category | Frequency (n) | Percentage (%) |
| --- | --- | --- | --- |
| pT Stage | pT2 | 42 | 34.7 |
|  | pT3 | 69 | 57.0 |
|  | pT4a | 5 | 4.1 |
|  | pT4b | 5 | 4.1 |
| pN Stage | pN0 | 70 | 57.9 |
|  | pN1 | 30 | 24.8 |
|  | pN2 | 18 | 14.9 |
|  | pN3 | 3 | 2.5 |
| Total Patients |  | 121 | 100.0 |

Postsurgical pathologic evaluation of the mass revealed that 76 (62.8%) patients had free surgical margins in the proximal, distal and radial margins, whereas 45 patients (37.2%) had positive radial margins. Among the 121 patients included in this study, 19 (15.7%) patients were documented to have experienced recurrence. Adjuvant chemotherapy after esophagectomy was given to 62 patients (51.2%). Among the 121 patients included in the study, 68 (56.2%) patients were on strict follow-up at their respective referral hospitals, and 53 (43.8%) patients were lost to follow-up.

The status of the operation was compared before and after surgery, as shown in Table 3. Among 121 patients who were confirmed as operable cases by preoperative CT TNM staging, 111 (93.9%) were actually operable, and 10 (6.1%) were inoperable, of which 5 were staged as T4 and the remaining 5 were staged as T3 on preoperative CT staging. Accordingly, preoperative CT staging had a 93.9% PPV for detecting operability, with 69.3% and 50% sensitivity and specificity, respectively.

**Table 3.**
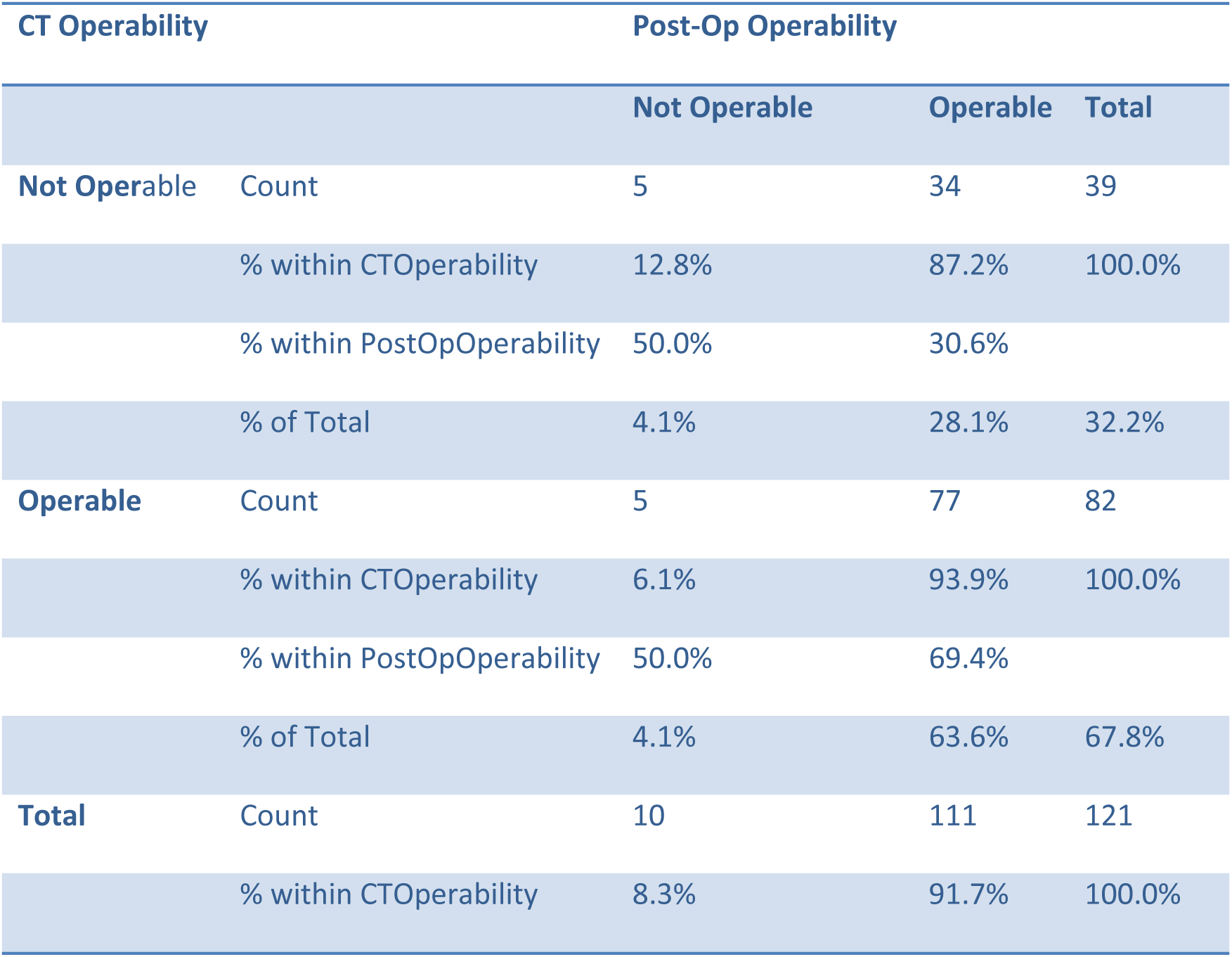
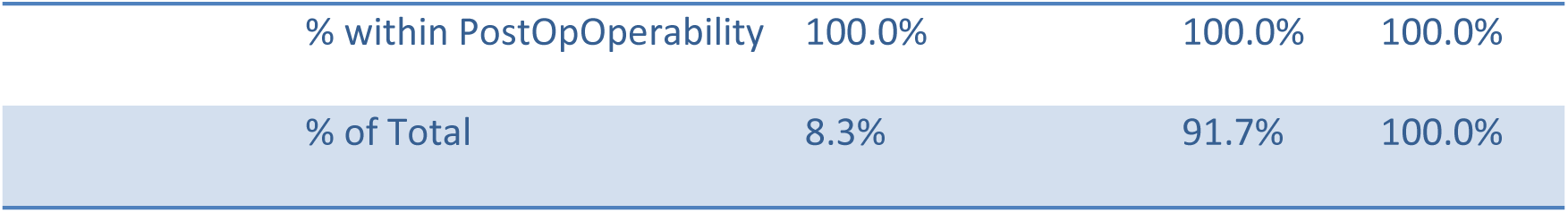
Comparison of surgical ability before and after surgery.

| CT Operability |  | Post-Op Operability |  |  |
| --- | --- | --- | --- | --- |
|  |  | Not Operable | Operable | Total |
| **Not Operable** | Count | 5 | 34 | 39 |
|  | % within CTOperability | 12.8% | 87.2% | 100.0% |
|  | % within PostOpOperability | 50.0% | 30.6% |  |
|  | % of Total | 4.1% | 28.1% | 32.2% |
| **Operable** | Count | 5 | 77 | 82 |
|  | % within CTOperability | 6.1% | 93.9% | 100.0% |
|  | % within PostOpOperability | 50.0% | 69.4% |  |
|  | % of Total | 4.1% | 63.6% | 67.8% |
| **Total** | Count | 10 | 111 | 121 |
|  | % within CTOperability | 8.3% | 91.7% | 100.0% |

|  |  |  |  |
| --- | --- | --- | --- |
| % within PostOpOperability | 100.0% | 100.0% | 100.0% |
| % of Total | 8.3% | 91.7% | 100.0% |

A comparison of the preoperative T stage and postsurgical pathological T stage of patients revealed that 66 (54.5%) patients were overstaged, 50 (41.3%) patients were correctly staged, and 5 (4.1%) patients were understaged, as shown in Table 4.

**Table 4.** Comparison between preoperative and postoperative T staging.

| CT Staging – | FREQUENCY | PERCENT |
| --- | --- | --- |
| Pathologic Staging |  |  |
| T3--PT2 | 32 | 26.4 |
| T3--PT3 | 45 | 37.2 |
| T3--PT4 | 5 | 4.1 |
| T4--PT2 | 10 | 8.3 |
| T4--PT3 | 24 | 19.8 |
| T4--PT4 | 5 | 4.1 |
| TOTAL | 121 | 100 |

According to the preoperative CT stage of T3, 32 (26.4%) patients had a pathological T2 stage, and 5 (4.1%) patients had a pathological T4 stage, as shown in Table 5. Only 45 (37.2%) patients had correctly staged T3 disease. Accordingly, the diagnostic accuracy of preoperative CT staging for T3 disease is 50%, with a sensitivity of 65.2%, specificity of 28.8%, PPV of 54.9% and NPV of 38.5%.

**Table 5.**
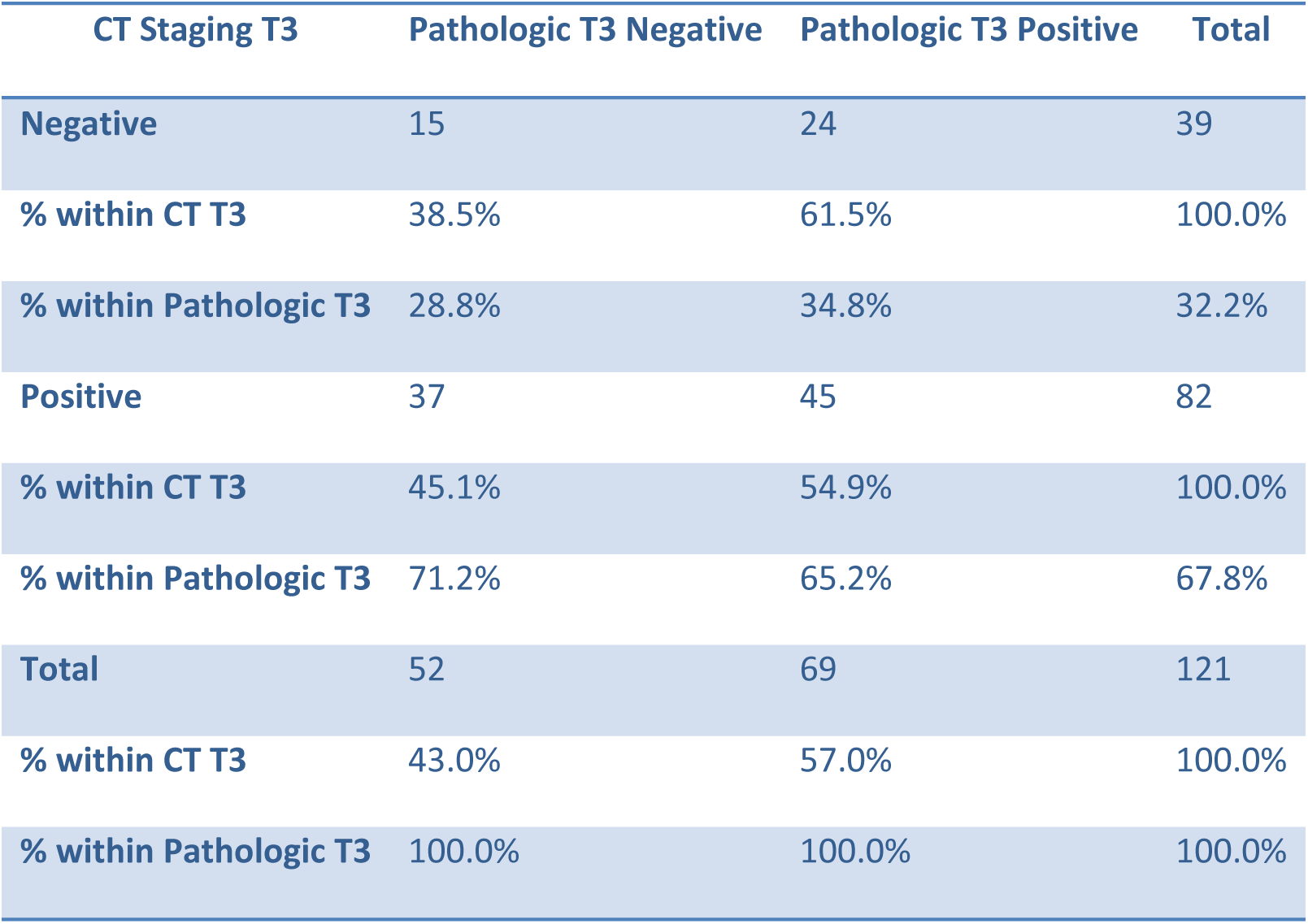
CT Staging T3 vs. Pathologic Staging T3 Crosstabulation.

From the preoperative CT stage of T4, 10 (8.3%) patients were found to have pathologic T2 disease, and 24 (19.8%) patients had pathologic T3 disease on postsurgical biopsy, as shown in Table 6. These patients were correctly staged at the preoperative T4 stage, which revealed that the diagnostic accuracy of preoperative CT staging for T4 esophageal disease was 67.8% in this study, with a sensitivity of 50%, specificity of 69.4%, PPV of 12.8% and NPV of 93.9%.

**Table 6.** CT Staging T4 vs. Pathologic Staging T4 Crosstabulation.

| CT Staging T4 | Pathologic T4 Negative | Pathologic T4 Positive | Total |
| --- | --- | --- | --- |
| Negative | 77 | 5 | 82 |
| % within CT T4 | 93.9% | 6.1% | 100.0% |
| % within Pathologic T4 | 69.4% | 50.0% | 67.8% |
| **Positive** | 34 | 5 | 39 |
| **% within CT T4** | 87.2% | 12.8% | 100.0% |
| **% within Pathologic T4** | 30.6% | 50.0% | 32.2% |
| **Total** | 111 | 10 | 121 |
| **% within CT T4** | 91.7% | 8.3% | 100.0% |
| **% within Pathologic T4** | 100.0% | 100.0% | 100.0% |

Seventeen (14%) patients were overstaged, 66 (54.5%) patients were correctly staged, and 38 (31.4%) patients were understaged. A total of 59 (48.7%) patients had correct staging as tumor-free nodes, 6 (4.9%) patients had correctly staged N1 esophageal cancer, 1 patient had correctly staged N2 disease, and none of the patients had a correct preoperative prediction for N3 disease, as shown in Table 7. The sensitivity and specificity of preoperative CT staging for N0 esophageal cancer patients are 84.3% and 31.4%, respectively. The sensitivity and specificity of preoperative CT staging for N1 disease are 90% and 20%, respectively, and those for N2 disease are 92.2% and 5.6%, respectively. In general, CT has a diagnostic accuracy, sensitivity, and specificity of 62%, 59% and 63%, respectively, for the presence of nodal disease. A comparison between CT and pathologic N staging for all patients can be found in Table 8.

**Table 7.** CT Detected Nodal Positivity vs Pathologic Nodal Positivity.

| **CT Node Positivity** | **Postop Node Negative** | **Postop Node Positive** | **Total** |
| --- | --- | --- | --- |
| **Negative** | 59 | 35 | 94 |
| **% within CT Negative** | 62.7% | 37.3% | 100.0% |
| **Positive** | 11 | 16 | 27 |
| **% within CT Positive** | 38.2% | 61.8% | 100.0% |
| **Total** | 70 | 51 | 121 |
| **% Total** | 100.0% | 100.0% | 100.0% |

**Table 8.**
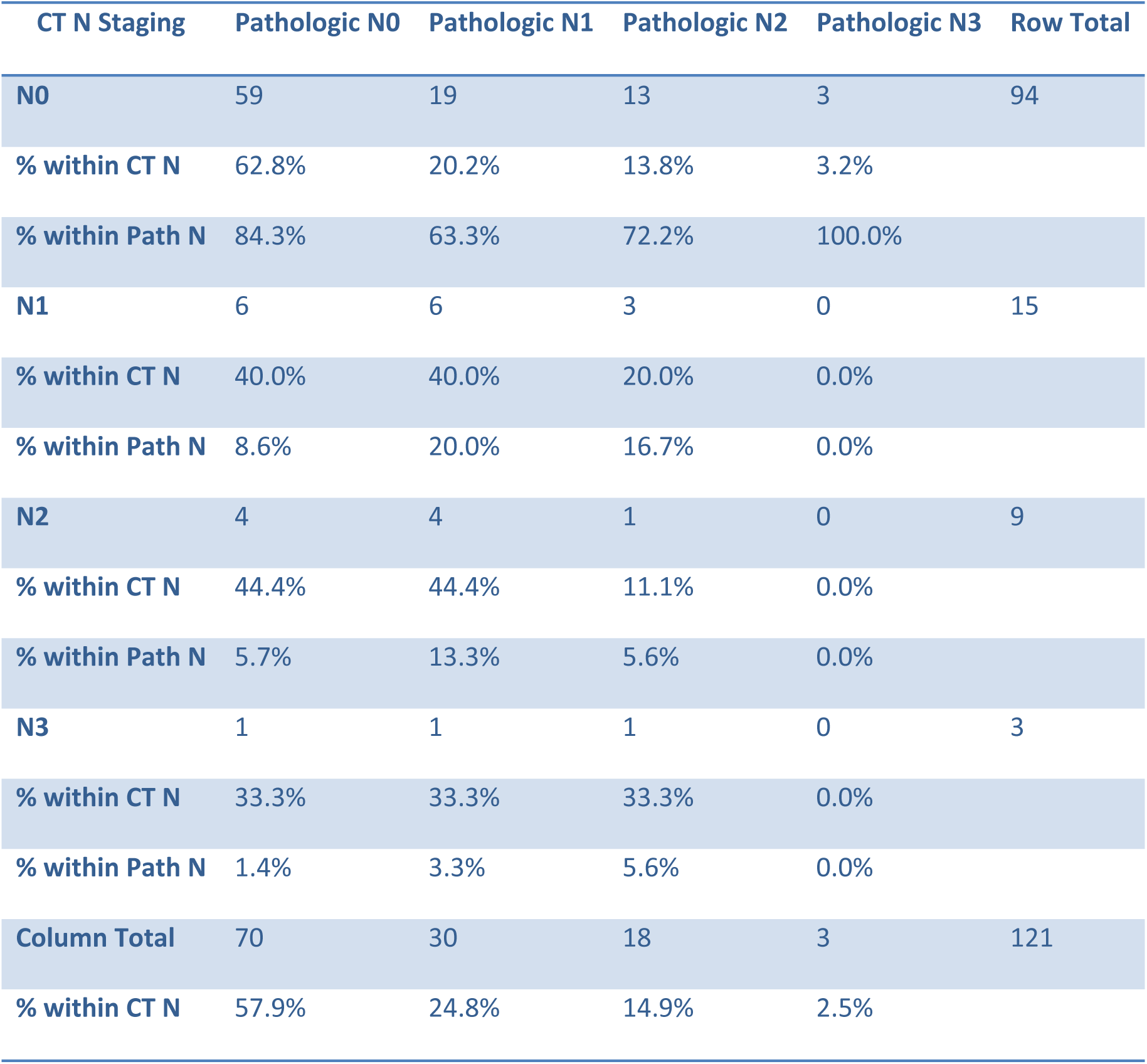
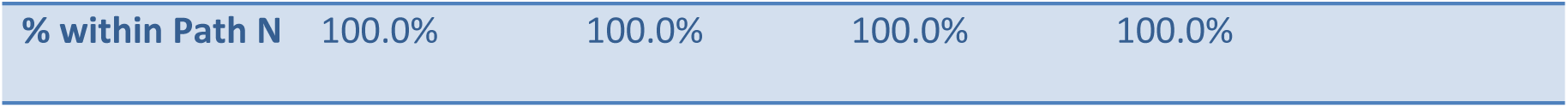
CT Staging N Disease vs Pathologic N Staging Cross-tabulation.

| **CT N Staging** | **Pathologic N0** | **Pathologic N1** | **Pathologic N2** | **Pathologic N3** | **Row Total** |
| --- | --- | --- | --- | --- | --- |
| **N0** | 59 | 19 | 13 | 3 | 94 |
| **% within CT N** | 62.8% | 20.2% | 13.8% | 3.2% |  |
| **% within Path N** | 84.3% | 63.3% | 72.2% | 100.0% |  |
| **N1** | 6 | 6 | 3 | 0 | 15 |
| **% within CT N** | 40.0% | 40.0% | 20.0% | 0.0% |  |
| **% within Path N** | 8.6% | 20.0% | 16.7% | 0.0% |  |
| **N2** | 4 | 4 | 1 | 0 | 9 |
| **% within CT N** | 44.4% | 44.4% | 11.1% | 0.0% |  |
| **% within Path N** | 5.7% | 13.3% | 5.6% | 0.0% |  |
| **N3** | 1 | 1 | 1 | 0 | 3 |
| **% within CT N** | 33.3% | 33.3% | 33.3% | 0.0% |  |
| **% within Path N** | 1.4% | 3.3% | 5.6% | 0.0% |  |
| **Column Total** | 70 | 30 | 18 | 3 | 121 |
| **% within CT N** | 57.9% | 24.8% | 14.9% | 2.5% |  |

|  |  |  |  |  |
| --- | --- | --- | --- | --- |
| % within Path N | 100.0% | 100.0% | 100.0% | 100.0% |

A total of 28 (23.1%) patients among 121 patients had correct T and N staging preoperatively, with a combined T and N staging sensitivity of 42.4% and specificity of 46.5%, as described in Table 9.

**Table 9.** Comparison of correct staging in both T and N stages.

| T_CorrectlyStaged | N_CorrectlyStaged: No | N_CorrectlyStaged: Yes | Total |
| --- | --- | --- | --- |
| No | 33 | 38 | 71 |
| % within T status | 46.5% | 53.5% | 100.0% |
| % within N status | 60.0% | 57.6% | 58.7% |
| Yes | 22 | 28 | 50 |
| % within T status | 44.0% | 56.0% | 100.0% |
| % within N status | 40.0% | 42.4% | 41.3% |
| Total | 55 | 66 | 121 |
| % within N status | 100.0% | 100.0% | 100.0% |

Overall, this study revealed that preoperative CT scan TNM staging had 82.6% specificity and 19.98% sensitivity for T staging and 59% sensitivity and 63% specificity for N staging, as shown in Table 10.

**Table 10.**
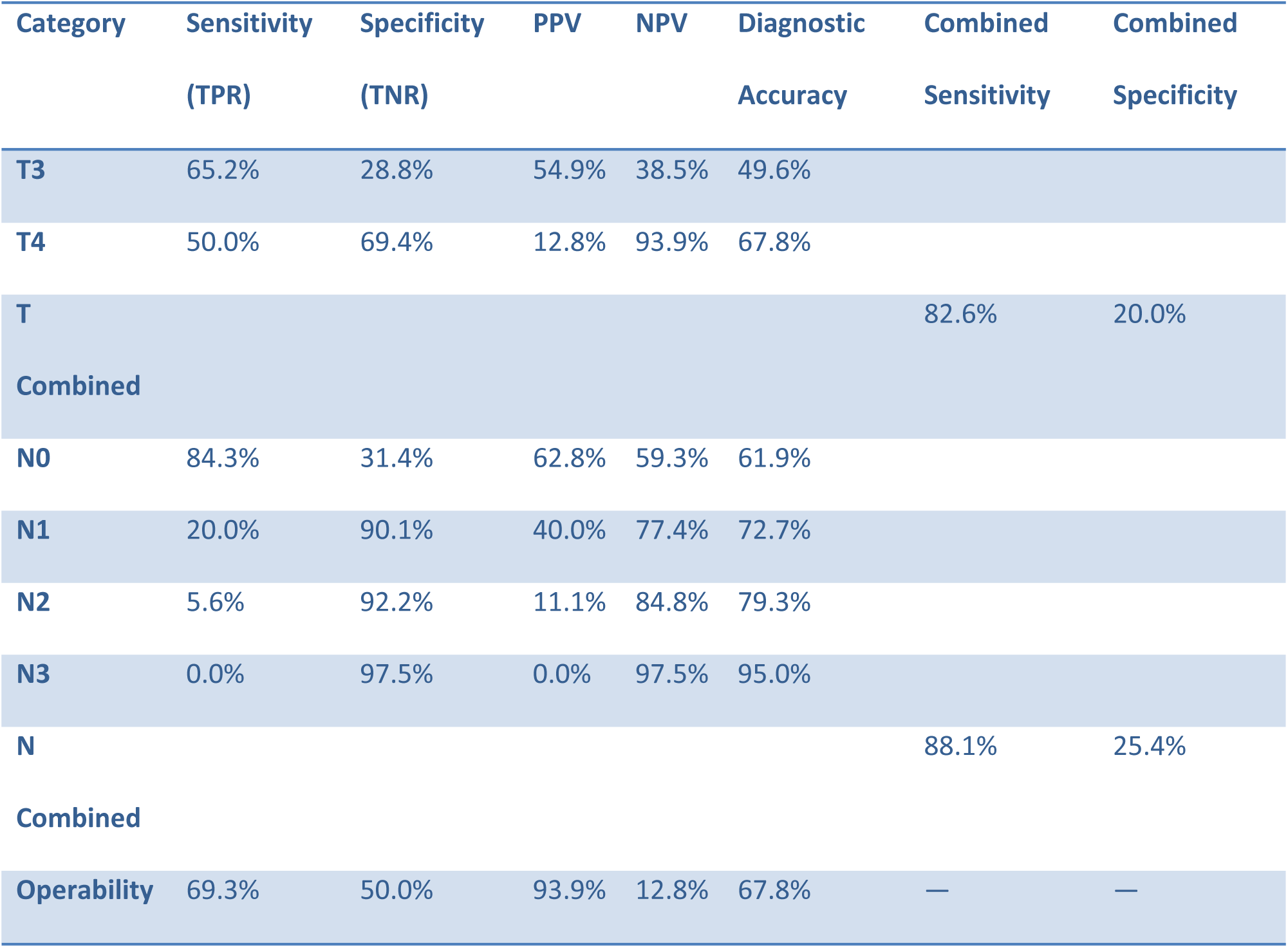
Measures of diagnostic test performance.

## Discussion

Accurate preoperative staging of esophageal cancer is essential for determining optimal treatment strategies. Our study assessed the diagnostic accuracy of preoperative CT TNM staging in predicting operability and compared it with postsurgical histopathological findings. The results demonstrated a tendency for CT to overstage T3 tumors while exhibiting moderate diagnostic accuracy for T4 tumors. These findings align with those of previous studies that highlighted the limitations of CT in differentiating tumor invasion depth and nodal involvement(2,9).

The overstaging of T3 tumors (54.5%) suggests that CT often misinterprets peritumoral inflammation or fibrosis as tumor invasion, a well-documented limitation of this imaging modality(10). However, the diagnostic accuracy of T4 staging was greater (67.8%), reflecting its better ability to detect clear signs of organ invasion. MRI has been proposed as a superior alternative for T-staging, as it offers better soft-tissue contrast and reduces motion artifacts(11). Recent studies indicate that 3T MRI outperforms CT in distinguishing early from advanced T stages, with a reported accuracy of up to 96%(12). While MRI is not widely used for routine staging, its improved diagnostic performance suggests potential applications in cases where CT findings are equivocal.

Our results indicated a significant tendency for understaging (31.4%) and overstaging (14%). The sensitivity of CT for detecting tumor-free nodes was high (84.3%), but the specificity was notably low (31.4%). These findings are consistent with previous reports that have questioned the reliability of CT for nodal assessment because of its dependence on size criteria rather than functional or metabolic characteristics (13). PET/CT radiomics has shown promise in improving N-staging accuracy by integrating metabolic and morphologic features, potentially addressing the limitations of size-based assessment alone (14). Studies have reported that compared with conventional CT, PET/CT-based radiomics achieves superior predictive performance (15).

The combined sensitivity and specificity of CT for T and N staging in our study were relatively low (42.4% and 46.5%, respectively), which suggests limitations in its ability to provide precise preoperative staging. Research has indicated that endoscopic ultrasound (EUS) may offer better accuracy for T staging, whereas PET/CT is advantageous for detecting nodal and distant metastases (16). Integrating these modalities into a multimodal staging approach may improve preoperative assessment and guide treatment planning more effectively.

Histopathologic analysis of our cohort confirmed squamous cell carcinoma (SCC) as the predominant histologic type (84.3%), which is consistent with epidemiological data from East African and Asian populations, where SCC is more prevalent than adenocarcinoma(17). The predominance of mid-esophageal tumors (82.8%) further supports regional trends in tumor location. Interestingly, our study revealed no significant correlation between staging accuracy and risk factors such as smoking or alcohol intake. This finding is consistent with prior literature suggesting that while these factors contribute to esophageal carcinogenesis, they do not directly influence imaging-based staging accuracy(11).

Surgical margin assessment revealed a negative margin rate of 62.8%, with 37.2% having positive radial margins. This finding highlights the challenge of achieving complete resection in esophageal cancer, particularly in advanced T stages. The recurrence rate of 15.7% within two years further emphasizes the importance of precise preoperative staging to ensure appropriate surgical candidacy and optimize long-term outcomes. Adjuvant chemotherapy was administered in 51.2% of the patients, reflecting the role of multimodal therapy in managing esophageal cancer.

## Conclusion

In conclusion, while CT remains a widely used imaging modality for preoperative staging, its limitations in T and N staging necessitate a multimodal approach incorporating MRI, EUS, and PET/CT when available. Future research should focus on the integration of advanced imaging techniques, including radiomics and artificial intelligence, to refine staging accuracy and improve patient outcomes.

## Data Availability

All relevant data are within the manuscript and its Supporting Information files.

## Acknowledgements

We would like to acknowledge Addis Ababa University, College of Health Sciences for providing the permission to conduct this study.

## Author Contributions

HT and MN conceptualized and wrote the proposal, ST collected the data, AM analyzed the data, MD wrote the discussion, MT compiled the manuscript and conducted the final review.

## Declaration of Generative AI and AI-assisted technologies in the writing process

During the preparation of this work, the author used Curie to improve language. After using this tool/service, the author reviewed and edited the content as needed and takes full responsibility for the content of the published article.

## Abbreviations

TASH: Tikur Anbessa Specialized Hospital
SCC: Squamous cell carcinoma
GEJ: Gastroesophageal Junction
CT: Computed Tomography
TNM: Tumor, Node, Metastasis (Staging system)
EUS: Endoscopic Ultrasound
PET: Positron Emission Tomography
MDT: Multidisciplinary Team
SPSS: Statistical Package for the Social Sciences
PPV: Positive Predictive Value
NPV: Negative Predictive Value
TPR: True positive rate (sensitivity)
TNR: True Negative Rate (Specificity)
THE: Transhiatal Esophagectomy
pTNM: Pathologic tumor, node, metastasis (staging system)
MRI: Magnetic Resonance Imaging
AJCC/UICC: American Joint Committee on Cancer/Union for International Cancer Control

